# Machine Learning-Driven Identification of Molecular Subgroups in Medulloblastoma via Gene Expression Profiling

**DOI:** 10.1101/2024.11.05.24316806

**Authors:** Hamdam Hourfar, Pegah Taklifi, Mahsa Razavi, Babak Khorsand

## Abstract

**Background:** Medulloblastoma (MB) is the most prevalent malignant brain tumor in children, characterized by substantial molecular heterogeneity across its subgroups. Accurate classification is pivotal for personalized treatment strategies and prognostic assessments.

**Procedure:** This study utilized machine learning (ML) techniques to analyze RNA sequencing data from 70 pediatric medulloblastoma samples. Five classifiers—K-nearest Neighbors (KNN), Decision Tree (DT), Support Vector Machine (SVM), Random Forest (RF), and Naive Bayes (NB)—were employed to predict molecular subgroups based on gene expression profiles. Feature selection identified gene subsets of varying sizes (750, 75, and 25 genes) to optimize classification accuracy.

**Results:** Initial analyses with the complete gene set lacked discriminative power. However, reduced feature sets significantly enhanced clustering and classification performance, particularly for Group 3 and Group 4 subgroups. The RF, KNN, and SVM classifiers consistently outperformed the DT and NB classifiers, achieving classification accuracies exceeding 90% in many scenarios, especially in Group 3 and Group 4.

**Conclusions:** This study highlights the efficacy of ML algorithms in classifying medulloblastoma subgroups using gene expression data. The integration of feature selection techniques substantially improves model performance, paving the way for enhanced personalized approaches in medulloblastoma management.

## 1. Introduction

Medulloblastoma (MB) is the most common malignant tumor of the central nervous system (CNS) in children, with an incidence of 5 cases per 1,000,000, making up approximately 20% of all pediatric brain tumors. In contrast, medulloblastoma is significantly rare in adults, with an incidence of 0.05 per 100,000 [1, 2]. Given that brain tumors are a leading cause of cancer-related morbidity and mortality in children, improving prognosis and developing more effective treatment strategies are essential for managing this condition [3].

While treatment advancements have improved survival rates for medulloblastoma over the years, they came at the cost of significant negative side effects. Standard treatment protocols typically involve a combination of surgery, radiation, and chemotherapy. However, these treatments carry the risk of long-term neurological damage, including cognitive impairments like memory loss [4]. For children under the age of three, radiation therapy is not considered safe due to its potential to severely disrupt brain development [4]. As a result, there is a critical need for the development of new therapies that target disease-causing cellular pathways while minimizing side effects and maximizing efficacy [5].

Recent technological advancements, such as Next Generation Sequencing (NGS) and DNA methylation profiling, have significantly deepened our understanding of the molecular heterogeneity of medulloblastoma. These advances offer promising opportunities for the development of more personalized and effective therapeutic interventions, and they represent a rapidly evolving area of research [3]. The discovery of distinct molecular subgroups of medulloblastoma began in 2002 when DNA microarray analysis of gene expression first revealed subgroup distinctions [6-9]. Since then, transcriptome-based studies have validated these findings [10], and it is widely accepted that medulloblastoma is comprised of at least four major molecular subgroups: WNT, SHH, Group 3, and Group 4, each with its own unique transcriptional, genetic, and clinical characteristics [4, 10].

WNT represents accounts for 10% of all medulloblastoma cases. It is associated with a highly favorable prognosis, with over 95% of patients achieving a 5-year overall survival (OS) rate. Metastasis is rare in this subgroup and the overexpression of β-catenin, which activates the WNT signaling pathway, is the main underlying molecular mechanism for this subgroup [4, 10].

The SHH subgroup comprises 25-30% of medulloblastomas. This subgroup has a 5-year OS rate of 70%, with prognosis varying by age group. Infants typically have a better outcome compared to older patients, where the prognosis is intermediate [4, 10]. SHH medulloblastomas are driven by the overexpression of the SHH signaling pathway. Additionally, around 30% of cases show abnormalities in the TP53 and PI3K pathways, leading to poorer prognosis and a reduced 5-year OS rate of 40% [1, 10]. Metastatic is relatively uncommon in the SHH subgroup [4].

Group 3 represents about 25% of all medulloblastomas. It is associated with the poorest prognosis among the subgroups, particularly in infants where the 5-year OS rate is just 45%. In older children, the OS improves slightly to 58%. The molecular mechanism underlying this subgroup remains unclear, which likely contributes to its poor prognosis [10]. However, TGF-β signaling is thought to play a role in the subgroup’s molecular mechanism [4]. Group3 also has the highest rate of metastasis at diagnosis and during recurrence [10].

The latest subgroup, Group 4 accounts for 35% of medulloblastomas with variable outcomes. Infants tend to fare worse, while older patients show 5-year OS rates ranging from 75 to 90% [10]. Like Group 3, the specific molecular drivers of Group 4 are not fully understood, but the nuclear factor NF-κB pathway has been implicated in its pathogenesis [4]. Although the metastatic rate in Group 4 is lower than in Group 3, frequent metastasis is still a concern [4, 10]. This substantial genetic heterogeneity observed among these molecular subgroups underscores the need for subgroup-specific, molecularly targeted treatments and more accurate prognostic tools [11]. Accurate molecular diagnosis of medulloblastoma subgroups based on genetic profiling is essential for delivering personalized therapies. To achieve this, novel technologies that enable precise diagnosis and prognosis prediction are critical for optimizing treatment strategies.

In this context, the integration of genomic data with artificial intelligence (AI) and machine learning (ML) approaches offers significant potential. AI and ML methods -both supervised and unsupervised-provide sophisticated statistical and computational tools for genomic classification and the identification of genetic risk variants [12]. These methods allow for precise prediction of disease subgroups and aid in developing personalized treatment plans.

In the present study, we employed ML algorithms to analyze RNA-seq gene expression data for the early diagnosis of medulloblastoma subgroups. Machine learning models are designed to learn complex patterns within gene expression data, distinguishing between the molecular subgroups pd medulloblastoma. Gene expression profiles were used as features and algorithms such as K-nearest Neighbour (KNN), Decision Tree (DT), Support Vector Machine (SVM), Naive Bayes (NB), and Random Forest (RF) were used as classifiers to predict the specific medulloblastoma subgroups.

## 2. Material and Methods

### 2.1 Dataset Collection and Clinical Data

In this study, we utilized a pediatric medulloblastoma dataset comprising 35,882 genes across 70 samples. The dataset was curated by integrating expression profiling data from two Gene Expression Omnibus (GEO) series: GSE143940 and GSE158413, which utilized high-throughput RNA sequencing (RNA-seq) technology. Both datasets employed the Illumina NextSeq500 sequencing platform, generating paired-end reads for accurate transcriptomic profiling [13]. This integration allowed us to obtain a comprehensive view of the gene expression landscape in pediatric medulloblastomas.

The 70 medulloblastoma cases in this study represent a wide range of pediatric ages, spanning from 0.3 to 18.3 years. Approximately 20.0% of the patients were aged three years or younger, while 80.0% were older than three years [13]. Additionally, there was an equal distribution of males and females, with a male-to-female ratio of 1:1 [10]. The molecular subgroup distribution of the patient samples included WNT (n=8, 11.4%), SHH (n=24, 34.3%), Group 3 (n=20, 28.6%), and Group 4 (n=18, 25.7%) [13]. Table 1 summarizes the clinical characteristics of the patient cohort.

**Table 1:**
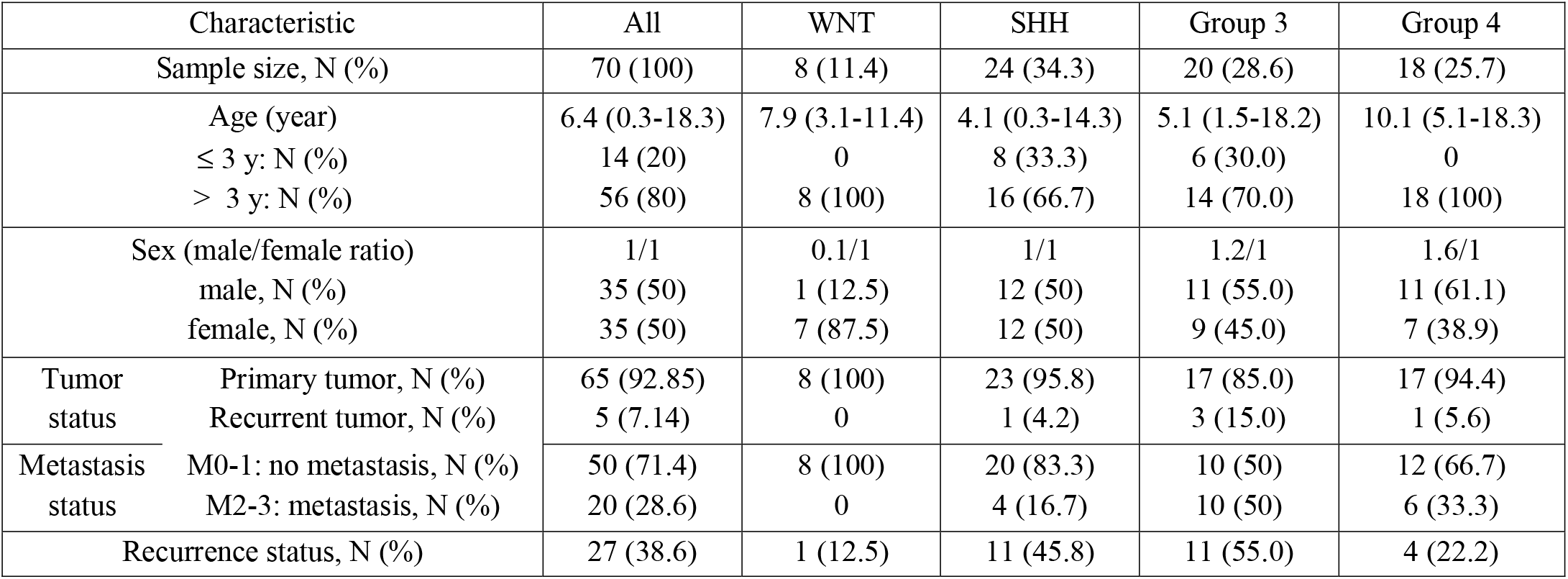
Clinical characteristics of pediatric medulloblastoma patients included in this study.

| Characteristic |  | All | WNT | SHH | Group 3 | Group 4 |
| --- | --- | --- | --- | --- | --- | --- |
| Sample size, N (%) |  | 70 (100) | 8 (11.4) | 24 (34.3) | 20 (28.6) | 18 (25.7) |
| Age (year) |  | 6.4 (0.3-18.3) | 7.9 (3.1-11.4) | 4.1 (0.3-14.3) | 5.1 (1.5-18.2) | 10.1 (5.1-18.3) |
| ≤ 3 y: N (%) |  | 14 (20) | 0 | 8 (33.3) | 6 (30.0) | 0 |
| > 3 y: N (%) |  | 56 (80) | 8 (100) | 16 (66.7) | 14 (70.0) | 18 (100) |
| Sex (male/female ratio) |  | 1/1 | 0.1/1 | 1/1 | 1.2/1 | 1.6/1 |
| male, N (%) |  | 35 (50) | 1 (12.5) | 12 (50) | 11 (55.0) | 11 (61.1) |
| female, N (%) |  | 35 (50) | 7 (87.5) | 12 (50) | 9 (45.0) | 7 (38.9) |
| Tumor status | Primary tumor, N (%) | 65 (92.85) | 8 (100) | 23 (95.8) | 17 (85.0) | 17 (94.4) |
|  | Recurrent tumor, N (%) | 5 (7.14) | 0 | 1 (4.2) | 3 (15.0) | 1 (5.6) |
| Metastasis status | M0-1: no metastasis, N (%) | 50 (71.4) | 8 (100) | 20 (83.3) | 10 (50) | 12 (66.7) |
|  | M2-3: metastasis, N (%) | 20 (28.6) | 0 | 4 (16.7) | 10 (50) | 6 (33.3) |
| Recurrence status, N (%) |  | 27 (38.6) | 1 (12.5) | 11 (45.8) | 11 (55.0) | 4 (22.2) |

### 2.2 Data Processing and Predicting Machine Learning Algorithms

This study aims to harness the predictive power of ML to classify medulloblastoma molecular subgroups of medulloblastoma according to key gene expression profile features. Processing data in machine learning is applied by five main types: supervised learning, unsupervised learning, semi-supervised learning, reinforcement learning, and deep learning [14, 15]. Supervised learning involves labeled input data to train models, while unsupervised learning clusters unlabeled data based on inherent similarities [14, 15]. Our primary objective for unsupervised learning was to cluster the gene expression profiles into logical groups based on shared features. K-Means, a widely used unsupervised clustering algorithm, was utilized to categorize the data into K clusters based on the similarity of gene expression profiles across samples [16]. For supervised learning, various machine learning algorithms and classifiers were applied to predict medulloblastoma subgroups, including K-Nearest Neighbors (KNN), Support Vector Machine (SVM), Decision Tree (DT), Random Forest (RF), and Naive Bayes (NB).

To determine which gene expression features contributed most to distinguishing medulloblastoma subgroups, we applied feature importance analysis. We identified three gene subsets based on feature rankings: 750, 75, and 25 genes, representing the most relevant genes across three different feature sets. In addition, we performed classification using the full dataset of 35,882 genes to compare performance across different feature subsets.

The data was split into training and testing sets using a stratified K-Fold cross-validation method (k=4) 75% for training and 25% for testing, ensuring the distribution of molecular subgroups was consistent across both subsets. This method helps to enhance model robustness and reduce bias, improving the model’s generalization to unseen data.

### 2.3 Evaluation Metrics

To evaluate the performance of each classification model, we used a range of commonly applied metrics, derived from the confusion matrix, which records the true positive (TP), true negative (TN), false positive (FP), and false negative (FN) predictions [17]. These metrics provide a detailed assessment of model accuracy and effectiveness in predicting medulloblastoma subgroups [17-20] :

- **Specificity** = TN / (TN + FP)
- **Sensitivity** (Recall) = TP / (TP + FN)
- **Precision** = TP / (TP + FP)
- **Negative Predictive Value (NPV)** = TN / (TN + FN)
- **Accuracy** = (TP + TN) / (TP + TN + FP + FN)
- **F1 Score** = 2TP / (2TP + FP + FN)

Additionally, we used the Matthews Correlation Coefficient (MCC), a robust metric for evaluating the performance of binary and multi-class classifiers, defined as:

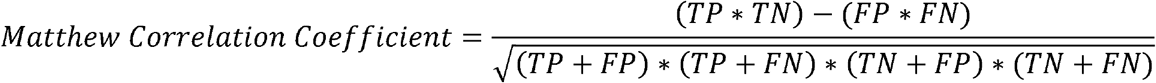

This suite of evaluation measures ensures a comprehensive assessment of the model’s prediction capabilities, allowing us to determine the most accurate and reliable algorithm for medulloblastoma subgroup classification.

## 3. Results

### 3.1 Clustering of Datasets

Initially, a clustering approach was applied to categorize data points into groups based on gene expression profiles of the RNA-seq dataset. The aim was to elucidate the clustering potential of the full gene expression profile (35882 genes) and its ability to differentiate between the intended molecular subgroups of medulloblastoma. However, the results indicated that the full gene expression profile lacked the discriminative power necessary for accurate classification of the distinct medulloblastoma subgroups. Moreover, there was inconsistency in data clustering, particularly for Group 3 and Group 4, across different iterations of the analysis (data not shown). These two subgroups, associated with poor and intermediate prognoses, respectively, are also known for their high metastasis rates (frequent in Group 3 and very frequent in Group 4) [4].

### 3.2 Clustering Based on Feature Selection

To improve the accuracy of clustering, a feature selectio strategy was employed, identifying gene subsets that demonstrated discriminative power. Three distinct subsets of genes were tested: 750, 75, and 25 genes with the highest feature scores. With the reduced feature sets, clustering performance improved significantly, leading to more precise classifications of three molecular subgroups: WNT, SHH, and Group 4. However, while Group 3 clustering was enhanced, some misclassifications still occurred (Figure 1). The enhanced clustering in the reduced feature sets underscores the importance of selecting key features for differentiating molecular subgroups of medulloblastoma, especially when dealing with a large dataset like RNA-seq profiles.

**Figure 1.**
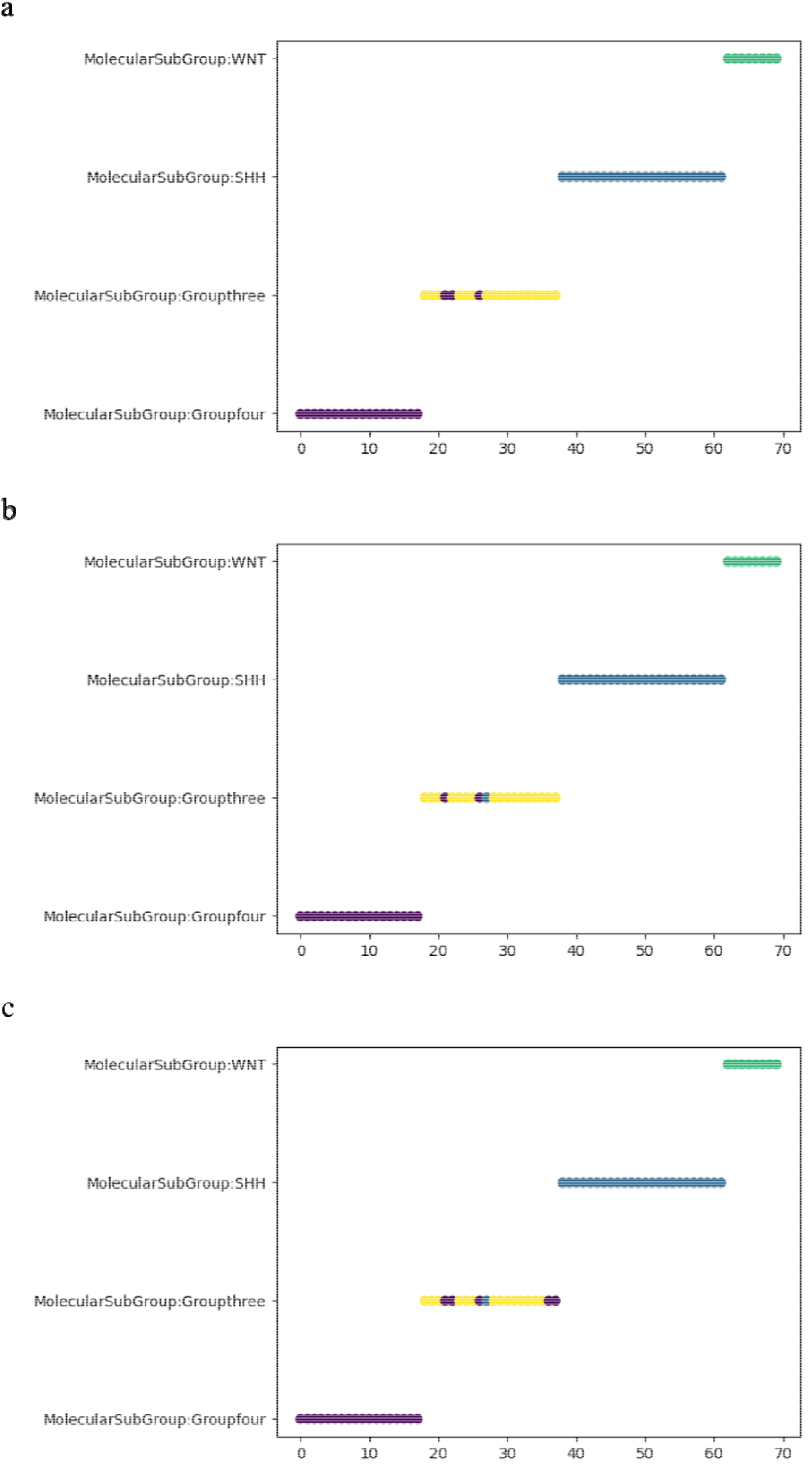
K-Means clustering plots using a) 750 genes, b) 75 genes, c) 25 genes.

### 3.3 Model Performance and Characteristics

To evaluate classification performance, the dataset was split into training, validation, and test sets, comprising 52 and 53 samples for training/validation, and 18 and 17 samples for testing. Multiple ML classifiers were applied, KNN, DT, SVM, RF, and NB. Seven performance metrics were calculated for each classifier. Four distinct gene sets were evaluated: 1) the entire gene expression profile (35, 882 genes), and those selected based on the highest feature importance score including 2) 750 selected genes, 3) 75 selected genes, and 4) 25 selected genes.

The classification results for each medulloblastoma molecular subgroup are summarized in Table S1. Notably, Group 3 and Group4 – subgroups typically associated with poor prognoses [4, 10]-showed significant performance improvements across the classifiers when reduced gene sets (759, 75, and 25 genes) were used. For instance, KNN, SVM, RF, and NB classifiers achieved performance metrics greater than 0.75, with many exceeding 0.85 and 0.9 across evaluation measures, particularly for Group3 and Group4. The DT classifier demonstrated more variable performance, with Group3 yielding superior results in the 750-gene feature set all compared to the entire 35,882-gene set, while results for Group4 were inconsistent across different metrics.

Across all feature subsets, KNN, SVM, RF, and NB classifiers consistently outperformed the DT classifier, especially classifying the SHH and WNT subgroups, where accuracy and other performance metrics frequently exceeded 0.9, with some models reaching perfect scores (1.0) in some cases 1). One exception was the NB classifier, which yielded a sensitivity of 0.75 for WNT classification in the 750-gene subset, likely due to the small sample size of this subgroup (n=8).

### 3.4 Performance Analysis of KNN Classifier

The performance of KNN across different feature subsets is illustrated in Figure S1. For Group 3, the 750-gene subset yielded a sensitivity of 0.85, an accuracy of 1, and an MCC of 0.90. Reducing the feature set to 25 genes still maintained high performance with a sensitivity of 0.75, an accuracy of 1, and an MCC of 0.82. Similar trends were observed for Group 4, with the 750-gene subset achieving perfect scores across sensitivity, accuracy, and MCC. These scores slightly decreased with the 25-gene subset, although sensitivity remained at 1. The KNN classifier consistently delivered excellent performance for the SHH and WNT subgroups, with near-perfect results, even in the 25-gene subset, highlighting the model’s robustness in classification.

### 3.5 Performance Analysis of Decision Tree (DT) Classifier

The DT classifier’s performance, as shown in Figure S2 revealed a more variable response compared to other algorithms. For most subgroups, the 75-gene feature subset underperformed relative to the 25- and 750-gene subsets, yielding results similar to those from the full 35,882-gene dataset (Figure S2). The SHH subgroup, in particular, demonstrated slightly better performance with the 25-gene subset compared to the 750-gene subset, while other subgroups showed nearly equivalent performance between these two feature sets.

### 3.6 Performance Analysis of SVM Classifier

The SVM classifier exhibited consistently strong performance across the three feature subsets (750, 75, and 25 genes), with a slight decline in accuracy for Group3 and SHH subgroups as the feature set size decreased (Figure S3). However, for Group4 and WNT, all three feature subsets resulted in identical scores, achieving perfect classification. The robustness of the SVM classifier, as evidenced by minimal performance degradation when reducing feature set size, suggests its high potential for accurate medulloblastoma subgroup classification.

### 3.7 Performance Analysis of Random Forest (RF) Classifier

The RF classifier achieved outstanding results across all gene feature subsets (Figure 2). It performed well with the 35,882-gene dataset, but performance improved even further when using the reduced subsets, with several subgroups achieving perfect classification scores. RF’s ability to generalize across varying feature set sizes underscores its reliability as a classification model for medulloblastoma subgroups.

**Figure 2.**
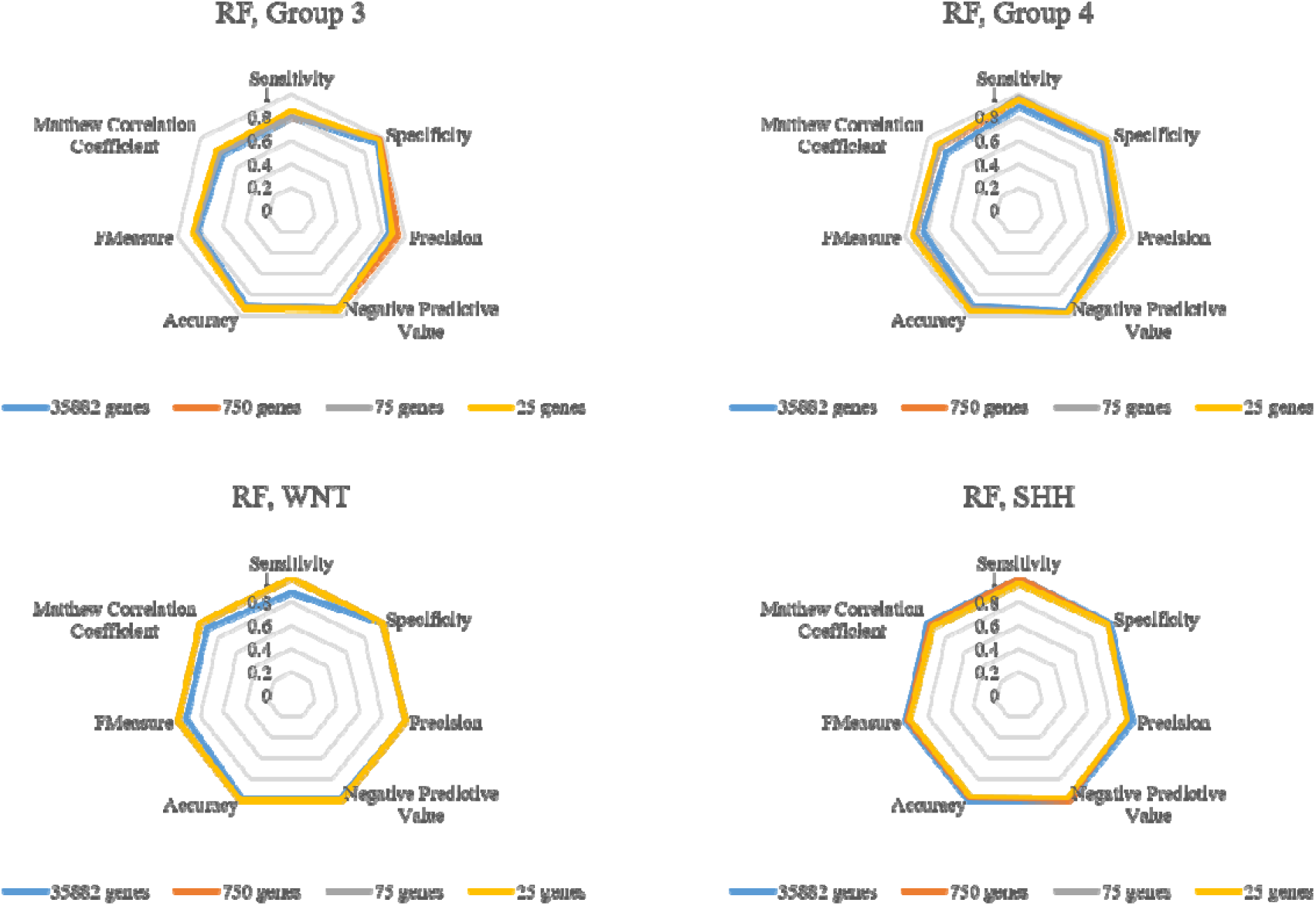
Evaluation metrics (sensitivity, specificity, precision, negative productive value, accuracy, F-measure, and Matthew correlation coefficient) for RF classifier across medulloblastoma subgroups.

### 3.8 Performance Analysis of Naive Bayes (NB) Classifier

The Naive Bayes (NB) classifier, applied to the 750-, 75-, and 25-gene subsets, demonstrated performance exceeding 0.8 across all metrics and subgroups, except sensitivity in the WNT subgroup using the 750-gene feature set, which yielded 0.75 (probably due to the small sample size for this subgroup (n=8)) (Figure S4). However, for the 25-gene feature set, the classifier showed strong performance across all metrics for Group3, Group 4, SHH, and WNT subgroups, achieving values exceeding 0.9 in most cases.

## 4. Discussion

This study aimed to identify and prioritize potential signature genes associated with the four molecular subgroups of medulloblastoma using machine learning approaches. Based on computational analysis, we selected three best feature subsets consisting of 750, 75, and 25 genes, representing the highest-scoring features. To uncover the genetic mechanisms underlying the classification of medulloblastoma into its molecular subgroups, we conducted gene ontology (GO) enrichment analysis on these feature subsets, in addition to identifying differentially expressed genes (DEGs) from the 750-gene subset through differential expression analysis (P value ≤ 0.05 & |log2FC| ≥1). The differential expression analysis was performed for three groupwise comparisons: 1) Group 3 versus Group 4, SHH, and WNT; 2) Group 4 versus SHH, and WNT; 3) SHH versus WNT. Notably, in some comparisons, few or no significant DEGs were identified, highlighting the complexity of the disease.

Gene ontology is a bioinformatics tool used to create computational representations of biological systems, which provides insights into gene product functions across species using standardized terms classified into three main categories: molecular function, cellular component, and biological process [21]. Molecular functions describe dynamic events (occurrents) such as catalytic activity, while cellular components represent physical structures (continuants) within the cell, like organelles or membranes [21, 22]. Biological processes refer to sequences of molecular events driven by multiple genes and proteins, facilitating cellular functions or organismal behavior [22]. The GO enrichment results analysis for the 750-gene feature subset and DEGs are provided in Supplementary Table S2.

The number of significant molecular function GO terms for the 750-, 75-, and 25-gene feature subsets, along with the DEGs and their respective up- and downregulated gene sets, were 95, 34, 27, 26, 30, and 7, respectively. Most of the enriched GO terms identified through DEG analysis were shared with at least one or more feature subsets, and five terms were found common across all three feature subsets. The 25- and 75-gene feature subsets shared three common GO terms with DEGs, while two terms were common between DEGs and the 25- and 750-gene feature subsets. Additionally, the 75- and 750-gene subsets shared two GO terms with the DEGs. Each feature subset represented a unique set of GO terms, with the 25-gene subset showing three, the 75-gene subset showing two, and the 750-gene subset showing seven GO terms shared with DEGs.

Interestingly, 48% of the GO terms in the 25-gene subset were shared with DEGs, and 7% of GO terms (2 of 27) were exclusively shared with downregulated genes. Similarly, 12 of 34 GO terms in the 75-gene feature subset (∼35%) overlapped with DEGs. This suggests that the 25-gene feature subset might have a slight advantage over the 75-gene subset, potentially due to the higher number of top-ranked genes, which could contribute to more accurate classification while eliminating less significant genes.

Notably, the two GO terms, Hyaluronic Acid Binding (GO:0005540) and Low-Density Lipoprotein Particle Receptor Binding (GO:0050750), were shared between the DEGs and the 25-gene feature subset. This indicates the potential relevance of these molecular functions in medulloblastoma pathogenesis and subgroup differentiation. As discussed in the introduction, medulloblastoma molecular subgroups exhibit distinct patterns of metastasis and chemoresistance [23]. Extracellular matrix (ECM)□associated genes and their diverse expression patterns have been shown to vary across the molecular subgroups of medulloblastoma, highlighting their potential role in tumor classification [23]. One such molecular function, Hyaluronic Acid Binding (GO:0005540), and its associated gene, *IMPG2*, which ranked as the third highest-scoring gene in our study, may serve as a valuable biomarker for distinguishing medulloblastoma subgroups.

Furthermore, cholesterol metabolism has been shown to play a crucial role in the SHH subgroup of medulloblastoma, particularly in tumor growth and maintenance of tumor-initiating cancer stem cells. Therapeutic strategies involving HDL nanoparticles have also been explored [24]. Cholesterol’s role in supporting the proliferation of medulloblastoma cells underscores its importance in subgroup classification [25]. PCSK9, a key regulator of cholesterol metabolism, emerged as one of the highest-scoring genes in our feature subset analysis. While PCSK9 primarily functions in cholesterol regulation in the liver, evidence suggests that it influences cholesterol homeostasis in the brain, particularly through its interaction with LDLR receptors, which mediate cholesterol transport into neurons [26].

PCSK9, under normal physiological conditions, cannot cross the blood-brain barrier (BBB), which restricts its regulatory function on cholesterol homeostasis to local expression within the central nervous system (CNS) [26]. This limitation in cholesterol regulation becomes particularly significant when considering medulloblastoma subgroups, as our study demonstrated markedly lower levels of *PCSK9* expression in Group 3 and Group 4 compared to the WNT subgroup. This reduced expression likely results in increased neuronal cholesterol within these groups, potentially promoting metastasis. Additionally, *PCSK9* was identified as one of the top 25 genes in our optimized feature subsets, highlighting its potential relevance in medulloblastoma subgroup classification.

In terms of gene expression dynamics, several genes exhibited their upregulation or downregulation when comparing different medulloblastoma subgroups, specifically among the 750 DEGs. For example, *NEUROG1*, the gene with the second-highest score in our analysis, presents differential expression between subgroups. It is downregulated in SHH versus WNT, but upregulated in Group 4 vs. SHH. This dual regulation underscores the complexity of *NEUROG1*’s role in subgroup-specific tumorigenesis. Previous studies have reported the variability in the expression of this gene among medulloblastoma patients, with some showing co-expression across certain groups, while others exhibit no co-expression [27]. Importantly, *NEUROG1* was absent in the normal cerebellum [27], further supporting its potential role as a subgroup-specific biomarker in medulloblastoma. While Northcott et al. [28] previously identified four distinct subgroups of medulloblastoma based on transcriptional profiles, this subgrouping according to expression profiles had been reported even earlier [29]. Furthermore, the overexpression of this neuronal survival gene, *NEUROG1*, in Group 4 medulloblastoma has been reported [30], consistent with our observations that it is a marker of this group.

Notably, *WIF1*, which had one of the highest scores in our study, exhibited significant downregulation in all non-WNT medulloblastoma subgroups compared to the WNT subgroup. This finding suggests a possible association between *WIF1* and this specific subgroup. A proposed mechanism suggests that WNT tumors secrete inhibitors of the Wnt pathway, such as WIF1 and Dickkopf-related protein 1 (DKK1), which could compromise vascular integrity by inhibiting Wnt signaling [31, 32]. This disruption may enhance the delivery of chemotherapy agents to tumor cells, providing an avenue for targeted treatment strategies specific to WNT medulloblastomas [33]. Moreover, WIF1 overexpression in the WNT subgroup, as observed in our results, could lead to changes in WIF1 secretion, thereby impacting the permeability of the BBB and facilitating drug delivery.

Similarly, members of the dickkop family, particularly *DKK4* and *DKK2*, have been previously reported as highly expressed in WNT medulloblastomas [34]. In line with this, our analysis also identified that *DKK4* and *DKK2* serve as potential biomarkers that can distinguish the WNT subgroup from others. Specifically, *DKK4*, which ranked among the top five features, was significantly downregulated across all non-WNT subgroups compared to the WNT subgroup. This consistent downregulation emphasized DKK4’s potential as a key marker for the WNT subgroup, as its reduced expression in other subgroups suggests a distinct molecular profile. Similarly, *DKK2* showed downregulation in Group3 and Group4 medulloblastomas when compared to the WNT subgroup in our study.

The WHO 2021 classification of medulloblastoma adds complexity to the landscape of predictive models, given its division into four principal molecular groups: WNT-activated, SHH-activated and *TP53*-wildtype, SHH-activated and *TP53*-mutant, and non-WNT/non-SHH (comprised of Group 3 and Group 4). Moreover, within these broader groups, additional subtypes have been identified, including four subtypes within the SHH group and eight subtypes for Group 3/4 [35, 36]. This intricate classification system can pose significant challenges for predictive models, especially those designed to handle complex genetic datasets. Despite this complexity, our optimized models have demonstrated strong performance, and their simplicity offers a practical advantage. This practical approach is particularly useful for situations where comprehensive molecular data may not be available or when simpler models are preferred for clinical implementation.

The WHO’s ongoing use of the four-subgroup classification of highlights the importance of differentiating between Group3 and Group4 medulloblastomas. Although these groups are often combined into the non-WNT/non-SHH category, they represent distinct subgroups with unique molecular characteristics and clinical behaviors. A precise distinction between Group 3 and Group 4 is critical for advancing therapeutic strategies. Each group’s genetic profile may reveal subgroup-specific therapeutic targets that could lead to more personalized and effective treatments for medulloblastoma patients. Identifying these differences could help tailor therapies to the biological makeup of each subgroup, improving treatment efficacy and patient outcomes.

In addition to the advantageous simplicity of our models, we used a comprehensive set of evaluation metrics, including sensitivity, specificity, precision, accuracy, negative predictive value, F-measure, and MCC, to assess the reliability of our classifiers. Notably, calculating these evaluation measures for each medulloblastoma subgroup offers a more refined assessment of how well each classifier performs in predictions.

While metrics like accuracy and F-measure are commonly used to assess model performance, they can be misleading when applied to imbalanced class datasets-a frequent challenge in cancer genomics [37, 38]. These metrics often fail to consider the ratio of positive to negative cases leading to poor performance when dealing with uneven class distributions. In contrast, metrics such as sensitivity, specificity, precision, and negative predictive value are more informative for such imbalanced datasets, as they provide insight into the model’s ability to correctly identify both positive and negative cases. By applying these measures to each medulloblastoma subgroup, we gained a clearer understanding of how well our models perform in identifying the specific characteristics of each group [39].

Furthermore, the inclusion of MCC as an evaluation metric adds another layer of robustness to our model assessments. MCC is particularly valuable in handling imbalanced datasets, as it takes into account true and false positives and negatives, providing a more balanced and reliable performance measure. MCC’s mathematical properties make it insensitive to class imbalances, ensuring that predictions remain accurate even when the dataset contains uneven representations of each subgroup [38]. By incorporating MCC into our evaluation, we were able to obtain more truthful and informative scores [38].

## 5. Conclusion

Accurate detection of medulloblastoma molecular subgroups plays a critical role in improving treatment decisions, as each subgroup exhibits unique genetic variations that can influence therapeutic outcomes. The correct classification of these subgroups is essential for selecting the most appropriate treatment strategies for patients. The results from our study indicate that the majority of our optimized models-especially those based on feature subsets-performed well, making them an effective tool for subgroup diagnosis and could aid in the development of more targeted therapeutic strategies, leading to better patient outcomes.

## Data Availability

All data produced in the present study are available upon reasonable request to the authors

## Table of Abbreviations

Abbreviation: Definition
MB: Medulloblastoma
CNS: Central Nervous System
NGS: Next Generation Sequencing
GEO: Gene Expression Omnibus
RNA-seq: RNA Sequencing
OS: Overall Survival
SHH: Sonic Hedgehog
WNT: Wingless/Integrated
TGF-β: Transforming Growth Factor Beta
SVM: Support Vector Machine
KNN: K-Nearest Neighbors
DT: Decision Tree
RF: Random Forest
NB: Naive Bayes
MCC: Matthews Correlation Coefficient
TP: True Positive
TN: True Negative
FP: False Positive
FN: False Negative
NPV: Negative Predictive Value
F1 Score: F1 Score
DEGs: Differentially Expressed Genes
FC: Fold Change
ECM: Extracellular Matrix
HDL: High-Density Lipoprotein
LDL: Low-Density Lipoprotein
PCSK9: Proprotein Convertase Subtilisin/Kexin Type 9
WIF1: Wnt Inhibitory Factor 1
DKK: Dickkopf (a Wnt Pathway Inhibitor)
WHO: World Health Organization

---

**Figure S1.**
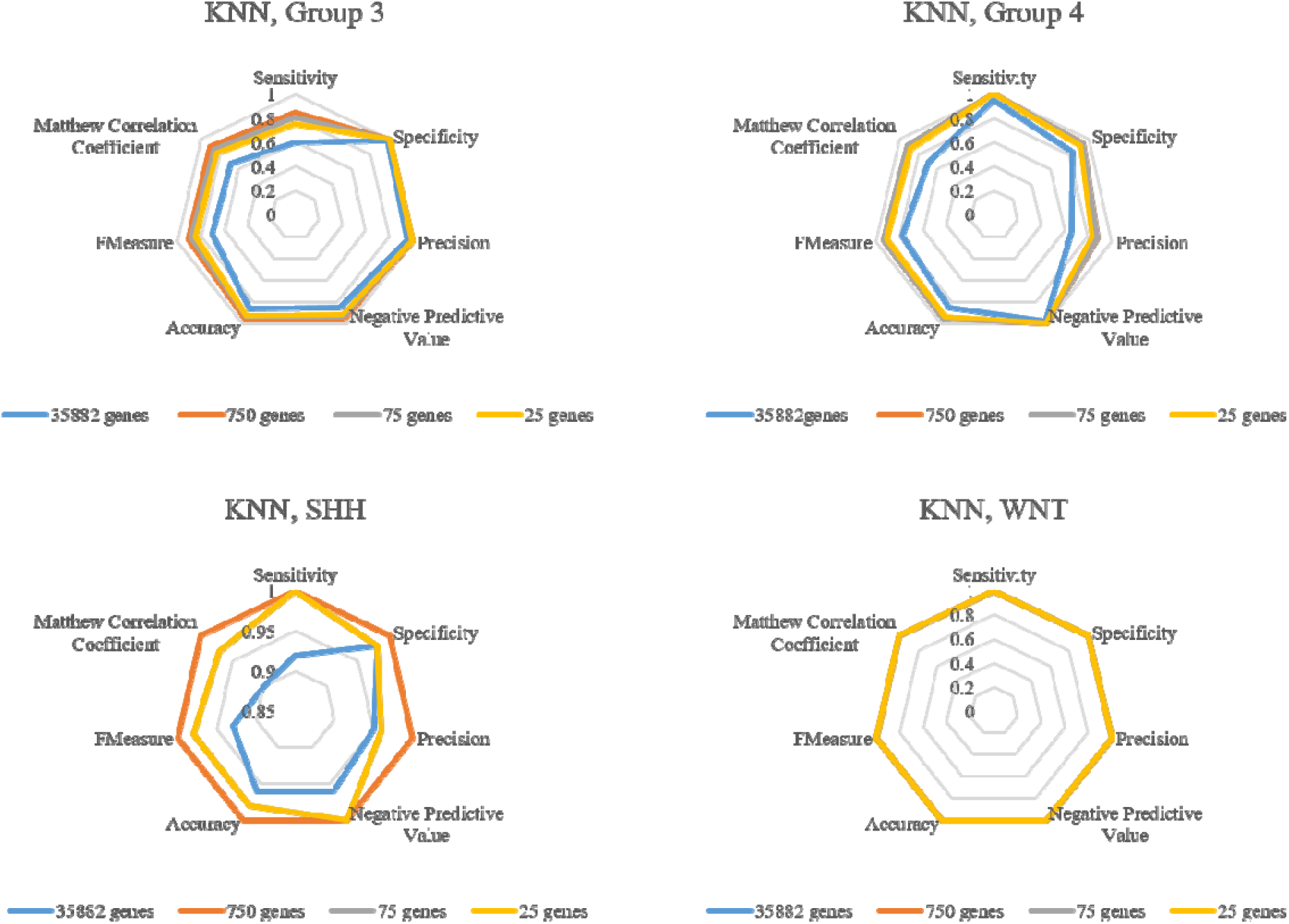
Evaluation metrics (sensitivity, specificity, precision, negative productive value, accuracy, F-measure, and Matthew correlation coefficient) for KNN classifier across medulloblastoma subgroups.

**Figure S2.**
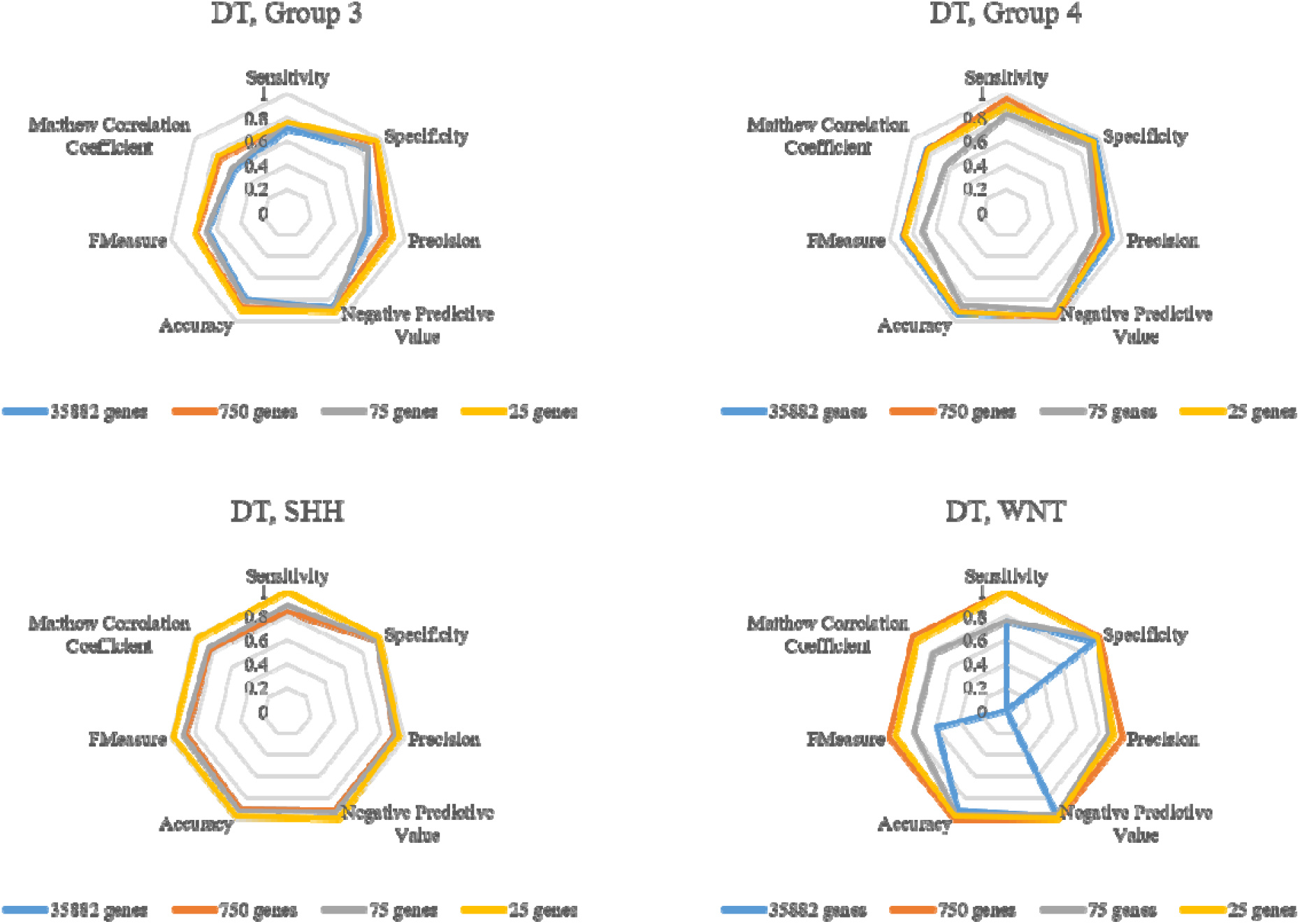
Evaluation metrics (sensitivity, specificity, precision, negative productive value, accuracy, F-measure, and Matthew correlation coefficient) for DT classifier across medulloblastoma subgroups.

**Figure S3.**
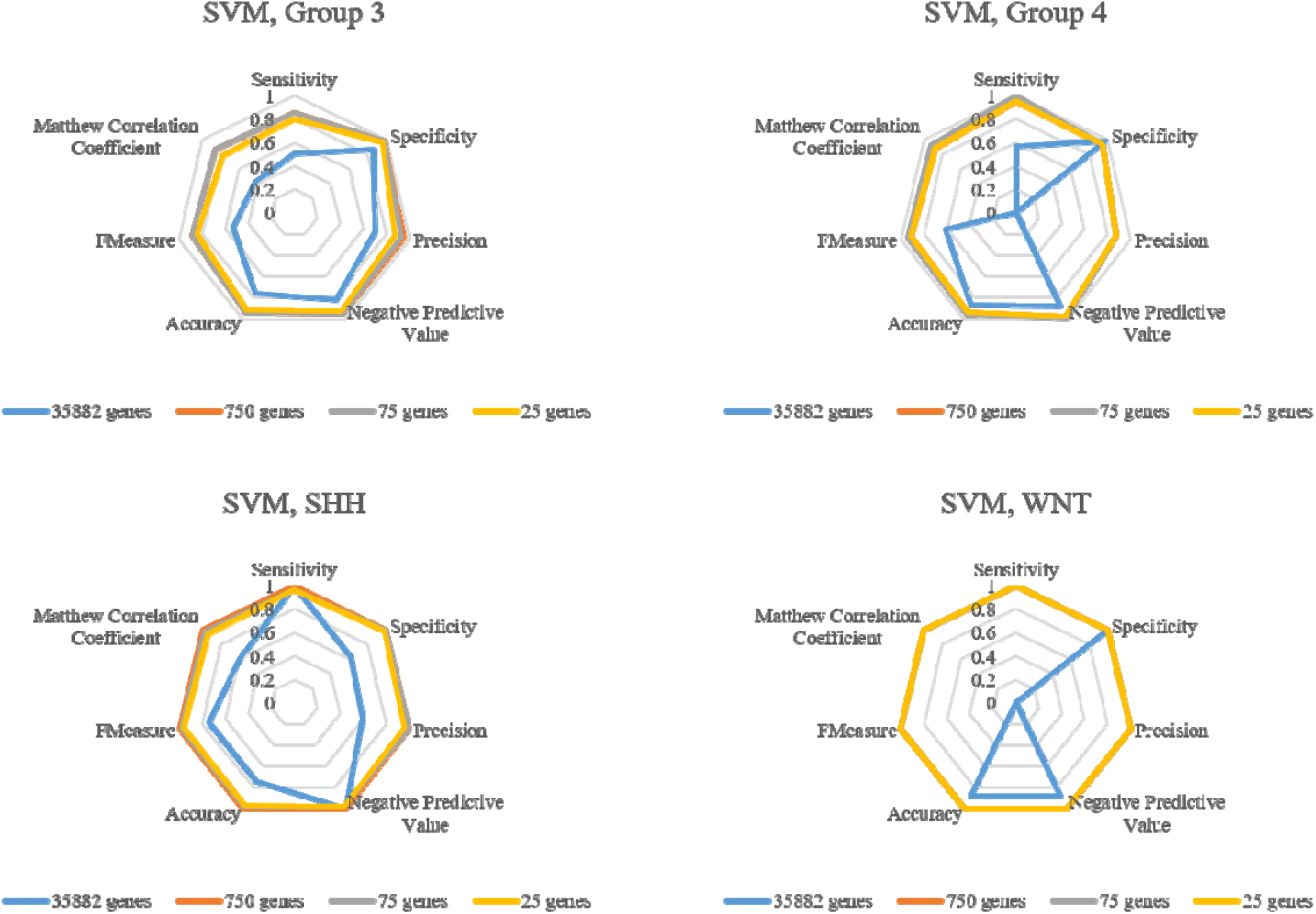
Evaluation metrics (sensitivity, specificity, precision, negative productive value, accuracy, F-measure, and Matthew correlation coefficient) for SVM classifier across medulloblastoma subgroups.

**Figure S4.**
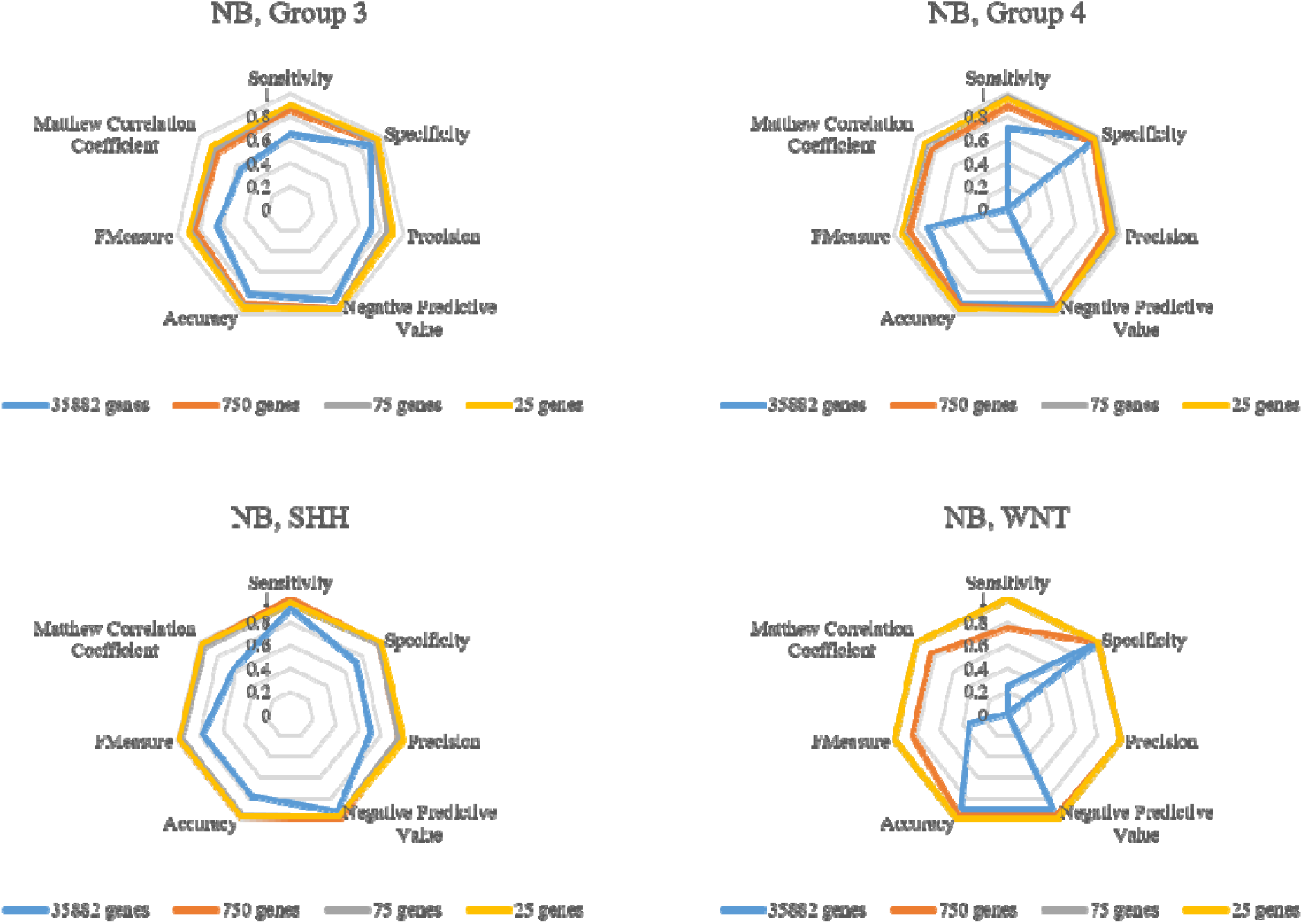
Evaluation metrics (sensitivity, specificity, precision, negative productive value, accuracy, F-measure, and Matthew correlation coefficient) for NB classifier across medulloblastoma subgroups.

